# Correlation between fat attenuation index and major adverse cardiovascular events: a systematic review and meta-analysis

**DOI:** 10.1101/2025.08.01.25332744

**Authors:** Yingzi Tan, Wang Yaojian, Zhao Boya, Yuerong Jiang, Keji Chen

**Author notes:** **Abbreviations and Acronyms** FAI, fat attenuation index; LAD, left anterior descending; LCX, left circumflex coronary artery; MACE, major adverse cardiovascular events; RCA, right coronary artery.

## Abstract

**Background:** The clinical predictive value of fat attenuation index (FAI) to major adverse cardiovascular events(MACE) remains uncertain.

**Objectives:** This study aimed to determine the relationship between FAI on coronary computed tomography angiography and MACE.

**Methods:** We searched four databases for cohort studies, case-control studies and cross-sectional studies on the relationship between FAI and MACE.We used RevMan5.4 for heterogeneity testing and statistical pooling. MACE was defined as the outcome measure, and publication bias was assessed using a funnel plot.

**Results:** 22 studies involving 10224 participants were enrolled. The results suggested that a significant correlation between adverse cardiovascular events and total FAI(FAI as a categorical variable: HR = 2.77, 95% CI = [2.22, 3.46], P < 0.00001, FAI as a continuous variable: HR = 1.15, 95% CI = [1.05, 1.26], P = 0.003), FAI of right coronary artery(RCA)branch(FAI as a categorical variable: HR = 2.10, 95% CI = [1.58, 2.79], P < 0.00001, FAI as a continuous variable: HR =1.06, 95% CI = [1.04, 1.08], P < 0.00001), FAI of left anterior descending (LAD) branch(FAI as a categorical variable: HR= 2.76, 95% CI= [1.93, 3.97], P < 0.00001, FAI as a continuous variable: HR= 1.09, 95% CI= [1.01,1.16], P < 0.00001), FAI of left circumflex coronary artery (LCX) branch(FAI as a categorical variable: HR=2.68, 95% CI=[1.24, 5.80], P=0.01, FAI as a continuous variable: HR=1.07, 95% CI = [1.05, 1.10], P < 0.00001),with individuals with elevated FAI levels having a significantly increased risk of developing adverse cardiovascular events. The funnel plot showed no publication bias.

**Conclusion:** As FAI levels increase, the risk of adverse cardiovascular events increases significantly. FAI may be a promising imaging indicator to predict MACE. (Correlation between fat attenuation index and major adverse cardiovascular events;CRD42022346488)

---

Cardiovascular disease (CVD) is the leading cause of death worldwide and a major threat to human health^[1,2]^. In the field of CVD prevention and treatment, accurate prediction of the risk of adverse events and guidance for early prevention in patients have always been important goals. In recent years, the rapid development of imaging technology has made the diagnosis and management of cardiovascular disease more convenient. CT angiography (CCTA) is now widely accepted as a first-line research tool for the detection of CAD, and the fat attenuation index (FAI) derived from coronary CCTA has received increasing attention as a novel, practical and non-invasive biomarker to reflect coronary artery inflammation in CVD risk assessment^[3]^.

Although there have been many studies on the relationship between FAI and cardiovascular adverse events, the conclusions of these studies are not entirely consistent. Some studies suggest a significant positive association between FAI and cardiovascular adverse events, with elevated FAI indicating a higher risk of cardiovascular adverse events^[4–5]^, while others suggest the opposite^[6]^. The inconsistency of research results not only confuses clinicians when using FAI for cardiovascular risk assessment, but also affects its promotion in clinical practice. Therefore, a meta-analysis of the relationship between FAI and cardiovascular adverse events is needed. This study aims to systematically evaluate the relationship between FAI and cardiovascular adverse events through meta-analysis to clarify its potential value in predicting and preventing cardiovascular disease. By comprehensively analysing data from multiple related studies, we hope to provide more accurate risk assessment tools for clinical practice and a scientific basis for future research directions.

## Methods

This meta-analysis strictly followed the PRISMA (Preferred Reporting Items for Systematic Reviews and Meta-Analyses) guidelines^[7]^.

### Published reports search

We searched PubMed, EMBASE, Web of Science and the Cochrane Library up to 3 March 2025. A combination of terms and free text terms were used for the search. The terms and key words are as follows: “Fat Attenuation Index”, “adverse cardiac events”, “myocardial infarction”, “stroke”, “heart failure”, “death, sudden, cardiac”, “MACE”, “myocardial revascularisation”, “severe arrhythmia”, “recurrent angina pectoris”. Using PubMed as an example, the specific search equation is shown in Table 1.

**Table 1.**
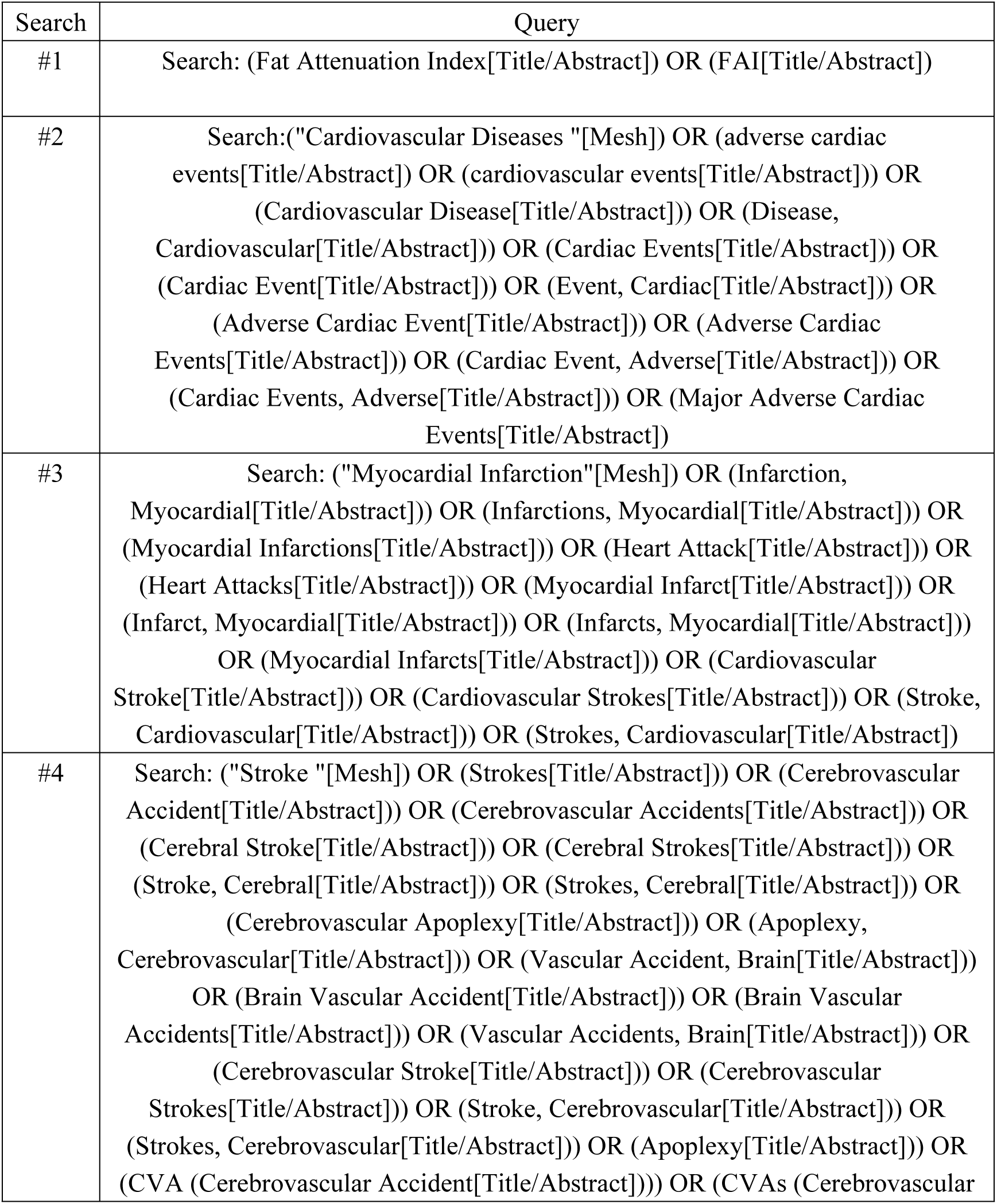
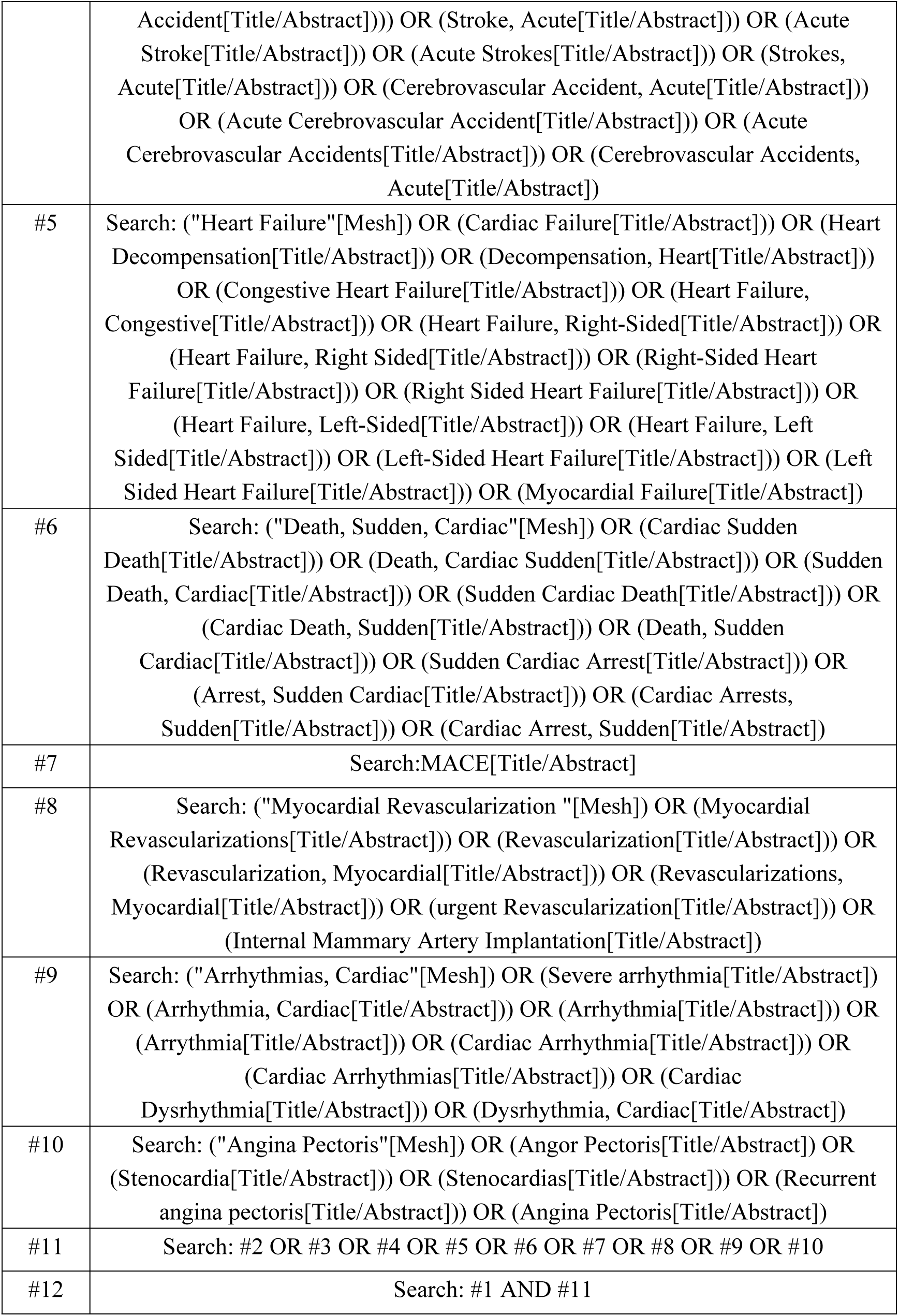
PubMed search strategy.

### Inclusion and Exclusion Criteria

Two authors (Y.T. and Y.J.) independently reviewed the literature based on strict inclusion and exclusion criteria. Inclusion criteria were developed as follows:(1)Population types: Patients diagnosed with or without CVD, regardless of age, sex, disease course, region, nationality, race, etc. (2) Types of outcomes: Correlation between outcome indicators FAI and MACE.(3)Types of study design: Prospective or retrospective observational study. (4) Exposure factor: FAI score. Exclusion criteria were as follows: (1) studies in which a hazard ratio (HR) could not be calculated; (2) duplicate studies or inability to obtain the full text of the literature; (3) literature that could not be extracted from the raw data.

### Data Extraction

The two researchers (Y.T. and Y.J.) worked independently to extract the following data from the included studies:(1) basic information such as first author’s name and year of publication;(2) characteristics of the research object such as sample size, types of study design, gender and age. Disagreements were resolved by discussion.

### Risk of bias assessment

According to the Newcastle-Ottawa Scale (NOS), the two researchers (Y.T. and Y.J.) independently assessed the risk of bias of the included studies. In the event of a dispute between the two researchers, the third party (K.C.) will mediate.

### Data analysis and synthesis

RevMan 5.4 software was used for data analysis. The hazard ratio (HR) was used as the effect indicator and its 95% confidence interval (CI) was provided. The heterogeneity of each group was assessed using the Q test and the I^2^ test. The fixed effect model would be chosen for analysis when P> 0.1 and I^2^ ≤50%, indicating that heterogeneity is modest. If P< 0.1 and I^2^ >50%, the heterogeneity is considered to be significant and further analysis of the sources of heterogeneity is required. After excluding the influence of significant clinical heterogeneity, the random-effects model would be selected for the trial. Use a funnel plot to assess publication bias for indicators with more than 10 included studies.

## Results

4 databases were searched for literature, yielding 802 documents: 361 in pubmed, 322 in Embase, 14 in Web of Science and 105 in the Cochrane Library. After importing all texts into NoteExpress and removing duplicate texts, 570 texts were obtained. After reading the title and abstract, 43 studies were obtained by removing the obviously ineligible ones. Then, 43 research articles were read in detail after being downloaded using different methods. 20 papers were excluded: data from 9 articles cannot be extracted^[4,5,8,9,10,11,12,13,14,15]^, the research indicators of 5 articles are radiomics^[16–20]^, the outcome measure of 5 articles is not MACE^[21–25]^ and 1 article is a review^[26]^.Finally, 22 high quality articles^[6,27–47]^ were screened. The process of document screening is shown in the following figure: (Fig. 1).

**Figure 1.**
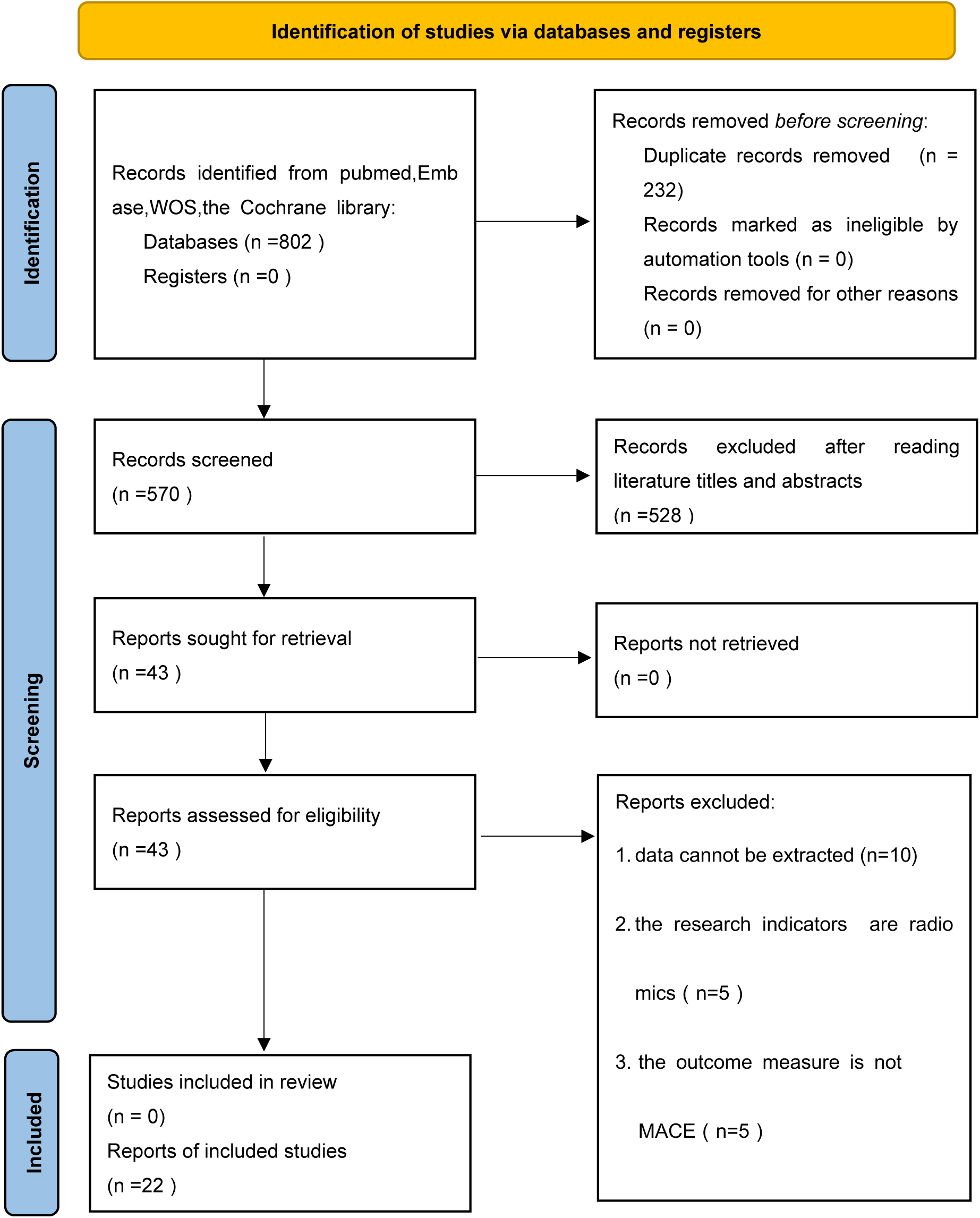
The flow diagram.

### Main characteristics of included studies

The 22 included articles^[6,27–47]^ were all published after 2021, ranging from 2021 to 2025. 13 studies^[30–36,41,43–47]^ are from China, 3 studies^[37,40,42]^ from Japan, 2 studies ^[38,39]^ from Italy, while the rest^[6,27,28,29]^ are from India, UK, USA and the Netherlands. A total of 10224 patients were included, with a maximum sample size of 3393 and a minimum sample size of 50. 17 studies^[6,28–30,32,35,36,38–47]^ used MACE as the outcome measure, 2 studies^[31,34]^ used acute coronary syndrome (ACS) as the outcome measure, 1 study^[37]^ used non-infarct-related territory unrecognised myocardial infarction (non-IR UMI) as the outcome measure, 1 study^[33]^ used unstable angina as the endpoint, 1 study^[27]^ used acute coronary events as the endpoint, 1 study^[40]^ used major adverse cardiac and cerebrovascular events (MACCE) as the endpoint. Basic information and characteristics of the included trials are shown in the table 2:

**Table 2.** General characteristics of the included review.

| Study | Country | Study type | Disease | Age | Gender (male/female) | Total | Publication year | Endpoint |
| --- | --- | --- | --- | --- | --- | --- | --- | --- |
| Biradar, B. <sup>[27]</sup> | India | Cohort study | CAD | 59.23±10.8 | 71/49 | 120 | 2025 | acute coronary events |
| Chan, K. <sup>[28]</sup> | UK | Cohort study | CAD | — | 1914/1479 | 3393 | 2024 | MACE |
| Chatterjee, D. <sup>[6]</sup> | USA | Cohort study | known or suspected CAD | — | — | 344 | 2021 | MACE |
| Coerkamp, C<sup>[29]</sup> | the Netherlands | Cohort study | suspected CAD | 62±7.7 | 20/30 | 50 | 2025 | MACE |
| Dai, X<sup>[30]</sup> | China | Cohort study | chest pain | 64.8±11.0 | 186/77 | 263 | 2022 | MACE |
| Huang, M<sup>[31]</sup> | China | Case control study | ACS | 59.3 ± 12.3 | 57/13 | 70 | 2023 | ACS |
| Huang, S. <sup>[32]</sup> | China | Cohort study | after aortic valve replacement | — | 87/52 | 139 | 2024 | MACE |
| Li, D.(1) <sup>[33]</sup> | China | Case control study | unstable angina | 64 ± 11.9 | 75/55 | 130 | 2025 | unstable angina |
| Li, D(2) <sup>[34]</sup> | China | Cohort study | stable angina pectoris | — | 156/122 | 278 | 2025 | ACS |
| Liu, M. <sup>[35]</sup> | China | Cohort study | type 2 diabetes mellitus (T2DM) | $61.79 \pm 9.86$ | 157/147 | 304 | 2024 | MACE |
| Luo, C<sup>[36]</sup> | China | Cohort study | CAD | — | 527/263 | 790 | 2024 | MACE |
| Matsuda, K<sup>[37]</sup> | Japan | Case control study | ACS | — | 121/37 | 158 | 2021 | non-IR UMI |
| Pergola, V<sup>[38]</sup> | Italy | Cohort study | no symptom or chest pain | $55.5 \pm 11.9$ | 237/134 | 371 | 2022 | MACE |
| Sansonetti, A. <sup>[39]</sup> | Italy | Cohort study | after heart transplant | $55.5 \pm 13$ | 63/38 | 101 | 2025 | MACE |
| Sayama, K. <sup>[40]</sup> | Japan | Case control study | takotsubo cardiomyopathy | $71 \pm 13$ | 10/42 | 52 | 2023 | MACCE |
| Sun, X. <sup>[41]</sup> | China | Cohort study | suspected CAD | $69.14 \pm 9.65$ | 153/107 | 260 | 2024 | MACE |
| Teng, Y. <sup>[42]</sup> | Japan | Cohort study | CCS | — | 145/36 | 181 | 2024 | MACE |
| Xie, Y. <sup>[43]</sup> | China | Cohort study | lung cancer | — | 345/352 | 697 | 2024 | MACE |
| Xu, Q. <sup>[44]</sup> | China | cross-sectional study | thoracic malignancies | — | 638/807 | 1543 | 2024 | MACE |
| Yu, Y. (1)<sup>[45]</sup> | China | Cohort study | suspected CAD | $60.6 \pm 9.9$ | 101/159 | 260 | 2024 | MACE |
| Yu, Y.(2) <sup>[46]</sup> | China | Cohort study | suspected CAD | 40.18 | 386/117 | 503 | 2025 | MACE |
| Zhang, X<sup>[47]</sup> | China | Cohort study | chest pain | $64.95 \pm 10.82$ | 139/78 | 217 | 2024 | MACE |

ACS = acute coronary syndrome; CAD = Coronary Artery Disease; CCS = Chronic Coronary Syndromes; MACCE = major adverse cardiac and cerebrovascular events; non-IR UMI = non-infarct-related territory unrecognised myocardial infarction

### Methodological evaluation of quality

In the risk of bias assessment, all included references received a score of 7 or higher, indicating high quality references. The results are shown in the following table: (Table 3-5).

**Table 3.** Quality Evaluation Form for Queue Research.

| Study | Year | Selectiveness | Comparability | Exposure | Total score |
| --- | --- | --- | --- | --- | --- |
| Biradar, B. | 2025 | 4 | 2 | 3 | 9 |
| Chan, K. | 2024 | 4 | 2 | 3 | 9 |
| Chatterjee, D. | 2021 | 4 | 0 | 3 | 7 |
| Dai, X | 2022 | 4 | 2 | 3 | 9 |
| Huang, S. | 2024 | 4 | 1 | 3 | 8 |
| Li, D(2) | 2025 | 4 | 2 | 3 | 9 |
| Liu, M. | 2024 | 4 | 2 | 3 | 9 |
| Luo, C | 2024 | 4 | 2 | 3 | 9 |
| Pergola, V | 2022 | 4 | 2 | 3 | 9 |
| Sansonetti, A. | 2025 | 4 | 2 | 3 | 9 |
| Sun, X. | 2024 | 4 | 2 | 3 | 9 |
| Teng, Y. | 2024 | 4 | 2 | 3 | 9 |
| Xie, Y. | 2024 | 4 | 2 | 3 | 9 |
| Yu, Y.(1) | 2024 | 4 | 2 | 3 | 9 |
| Yu, Y.(2) | 2025 | 4 | 2 | 3 | 9 |
| Zhang, X | 2024 | 4 | 2 | 3 | 9 |
| Coerkamp, C | 2025 | 4 | 2 | 3 | 9 |

**Table 4.** Quality Evaluation Form for Case Control Studies.

| Study | Study | Selectiveness | Comparability | Exposure | Total score |
| --- | --- | --- | --- | --- | --- |
| Huang, M | 2023 | 4 | 2 | 3 | 9 |
| Sayama, K. | 2023 | 4 | 2 | 3 | 9 |
| Li, D.(1) | 2025 | 4 | 2 | 3 | 9 |
| Matsuda, K | 2021 | 4 | 2 | 3 | 9 |

**Table 5.** Quality Evaluation Form for cross-sectional study.

| Study | Study | Selectiveness | Comparability | Exposure | Total score |
| --- | --- | --- | --- | --- | --- |
| Xu, Q. | 2024 | 4 | 2 | 3 | 9 |

### Meta-analysis

A total of 18 studies[27,28,29,30,31,33,34,35,36,37,38,39,40,41,42,45,46,47] focused on the relationship between total FAI and MACE. Depending on the statistical methods used for FAI, it can be divided into two categories: FAI as a categorical variable (12 studies [29,33,34,35,37,38,39,40,42,45,46,47]) and FAI as a continuous variable (7 studies [27,28,30,31,36,40,41]). 1 study^[40]^ used both categorical and continuous variables for FAI.

In statistical models, when FAI is used as a continuous variable, the hazard ratio (HR) represents the proportion of increased risk of MACE for each HU unit increase. In statistical models, when FAI is used as a categorical variable, HR represents the risk ratio of MACE for a given category (such as the high FAI group) relative to the reference category (such as the low FAI group).

### Correlation between total FAI and MACE (FAI as a categorical variable)

A total of 12 studies^[29,33,34,35,37,38,39,40,42,45,46,47]^ evaluated the correlation between FAI and MACE as a categorical variable. The heterogeneity test showed chi^2^=11.65, DF=11(P=0.39) and I^2^=6%, indicating a low level of heterogeneity. We used fixed model meta-analysis. When FAI is used as a categorical variable, the risk of adverse cardiovascular events in the high FAI group is 2.77 times higher than that in the low FAI group (HR=2.77,95%Cl=[2.22,3.46],P<0.00001), according to the statistically significant results.(Figure 2) When individual studies were removed one by one for sensitivity analysis, no significant differences in the results of individual studies were observed (I^2^ was 0% ∼18%, HR was 2.58∼3.00), indicating that the results of this meta-analysis are relatively stable and reliable.

**Figure 2.**
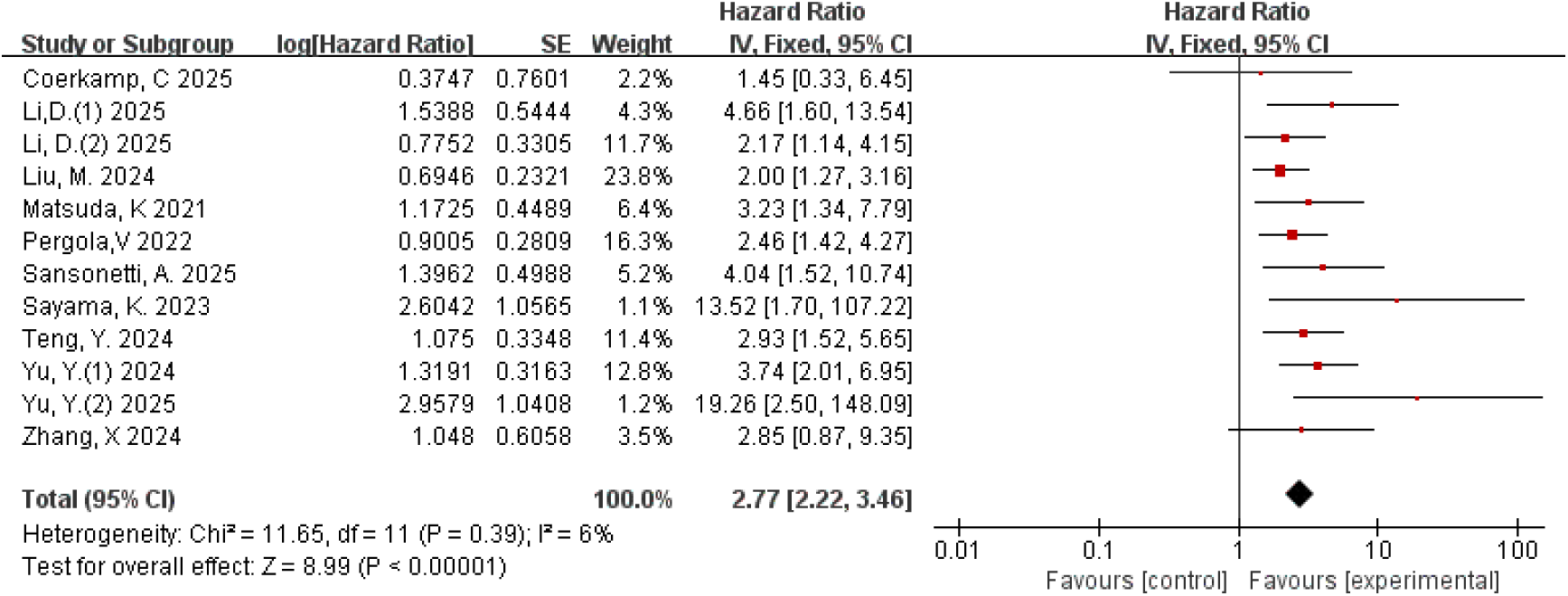
Forest plots of the relationship between total FAI and MACE(FAI as a categorical variable)

### The publication bias of the correlation between total FAI and MACE

The distribution of the scatter points in the funnel plot is roughly concentrated, and each scatter point is symmetrical along the two sides of the dotted line, indicating that the publication bias is minimal and the results are trustworthy. The funnel plot as figure 3 shown below:

**Figure 3.**
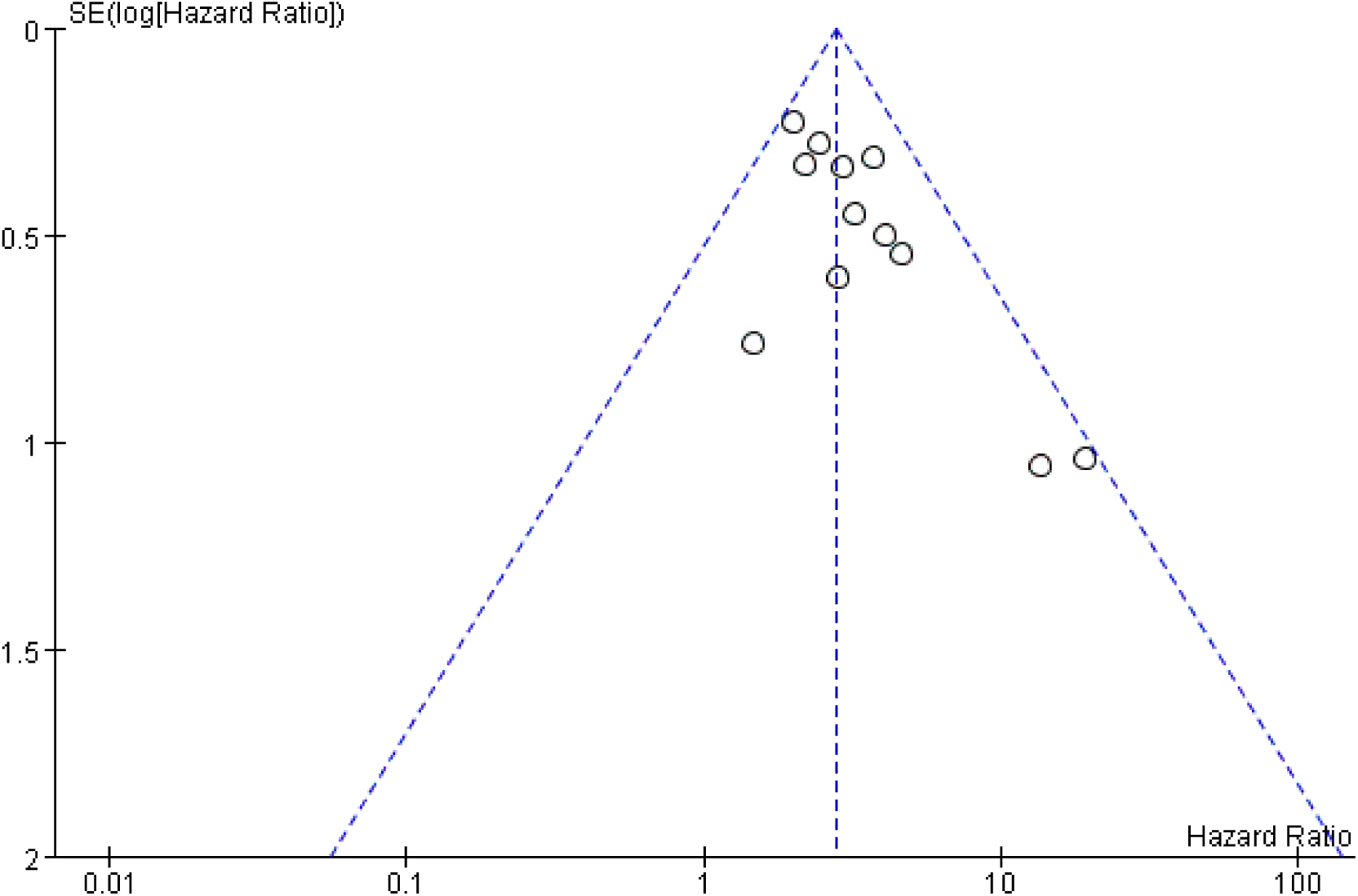
Funnel plot of publication bias.

### Correlation between total FAI and MACE (FAI as a continuous variable)

7 studies^[27,28,30,31,36,40,41]^ evaluated the correlation between FAI and MACE as a continuous variable. The heterogeneity test showed chi^2^ = 87.52, DF = 6(P<0.00001) and I^2^ = 93%, indicating a high degree of heterogeneity. We used random model meta-analysis. The overall effect size is as shown in the figure: HR = 1.15, 95% CI = [1.05, 1.26], P = 0.003. According to the statistically significant results, when FAI is used as a continuous variable, the risk of MACE increases by 15% for each additional HU unit. The forest plots are as figure 4:

**Figure 4.**
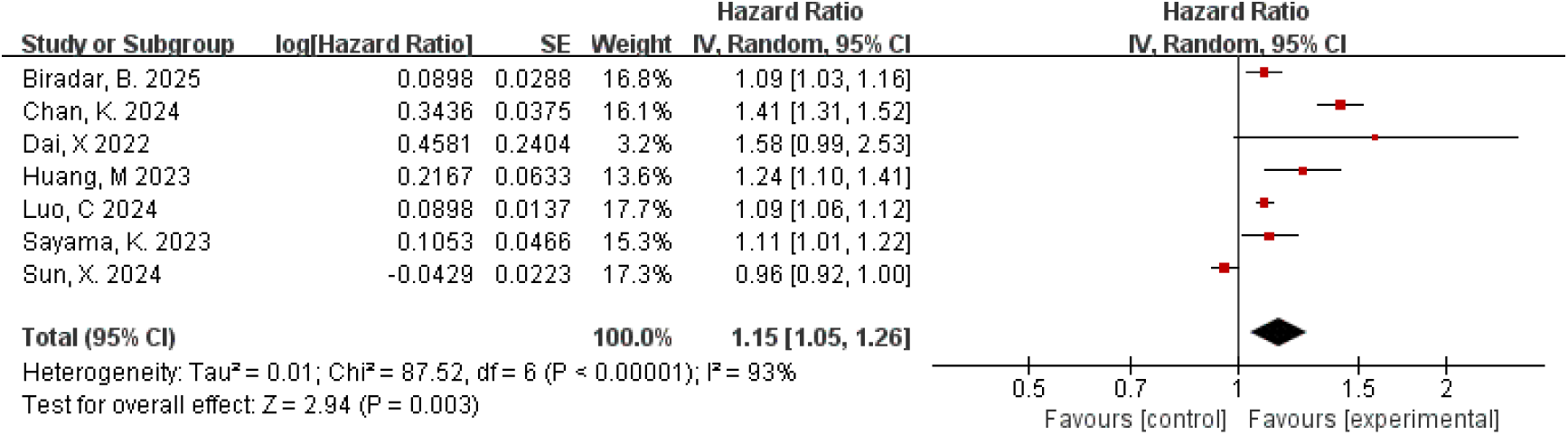
Forest plots of the relationship between total FAI and MACE(FAI as a continuous variable)

After excluding 2 studies^[28,41]^ with no statistically significant differences in data, I^2^=35%.The heterogeneity is small and a fixed model meta-analysis was performed with a combined effect size of HR=1.10, 95% CI=[1.08, 1.13], P<0.00001.

Reasons for heterogeneity could be: 1. The results of 1 study^[41]^ are different from those of other studies, which may be one of the reasons for heterogeneity; 2. The confidence interval range of each study is relatively small; 3. The statistical method of 1 study^[30]^ is different from other studies, and its HR in this study is the effect of increasing the proportion of MACE per 10 HU increase, while in other studies it is the effect of increasing the proportion of MACE per 1 HU increase; 4. The different studies come from different research centres, with different instruments, measurement methods and statistical methods, which may cause deviations in the calculation results. The results of this meta-analysis are based on the heterogeneity caused by the experimental results, which have statistical differences and indicate a strong correlation between total FAI and MACE. See as figure 5.

**Figure 5.**
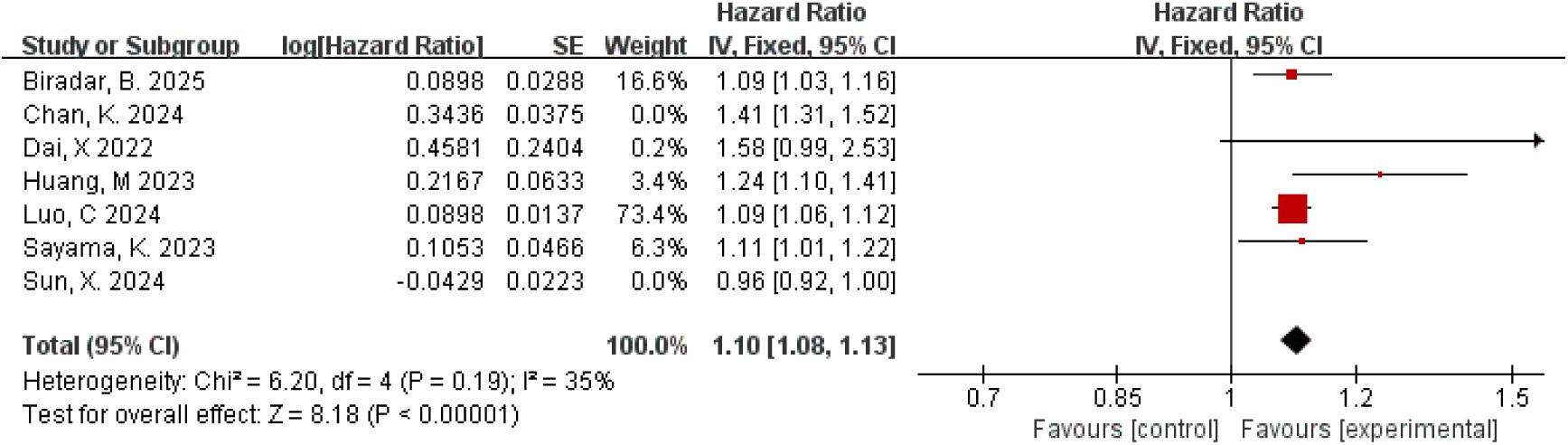
sensitivity analysis of the relationship between total FAI and MACE(FAI as a continuous variable)

### Correlation between FAI of RCA branch and MACE (FAI as a categorical variable)

The data is derived from 4^[32,35,43,44]^literature. The heterogeneity test resulted in chi^2^ = 5.24, DF =3(P =0.15), and I^2^ =43%, indicating a low level of heterogeneity. We employed fixed model meta analysis. The overall effect amount is as given in the figure: HR = 2.10, 95% CI = [1.58, 2.79], P < 0.00001. There is a significant correlation between FAI of RCA branch and MACE, according to the statistically significant results.(Figure 6)

**Figure 6.**
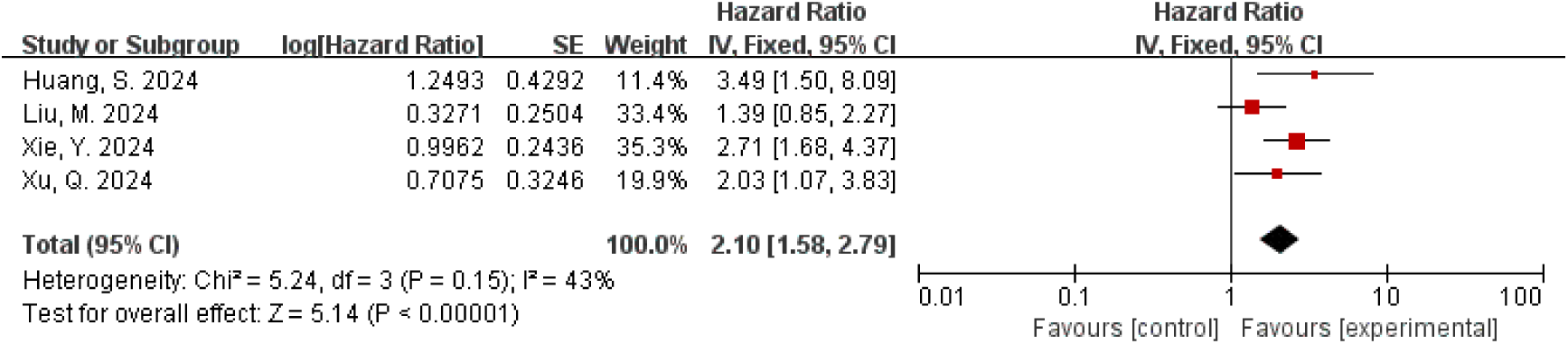
Forest plots of the relationship between FAI of RCA branch and MACE(FAI as a categorical variable)

### Correlation between FAI of RCA branch and MACE (FAI as a continuous variable)

The data is derived from 3^[6,36,40]^ literature. The heterogeneity test resulted in chi^2^=0.32, DF=2(P=0.85), and I^2^=0%, indicating a low level of heterogeneity. We employed fixed model meta analysis. The overall effect amount is as given in the figure: HR =1.06, 95% CI = [1.04, 1.08], P < 0.00001. There is a significant correlation between FAI of RCA branch and MACE, according to the statistically significant results.(Figure 7)

**Figure 7.**
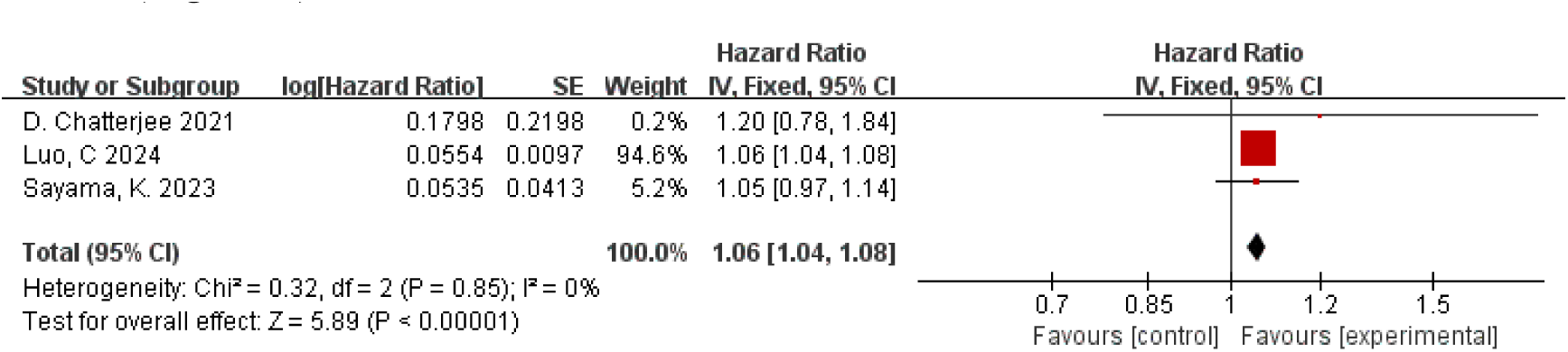
Forest plots of the relationship between FAI of RCA branch and MACE(FAI as a continuous variable)

### Correlation between FAI of LAD branch and MACE (FAI as a categorical variable)

The data is derived from 3^[35,43,44]^ literature. The heterogeneity test resulted in chi^2^=3.29, DF=2(P= 0.19), and I^2^ = 39%, indicating a low level of heterogeneity. We employed fixed model meta analysis. The overall effect amount is as given in the figure: HR= 2.76, 95% CI= [1.93, 3.97], P < 0.00001. There is a significant correlation between FAI of LAD branch and adverse cardiovascular events, according to the statistically significant results.(Figure 8)

**Figure 8.**
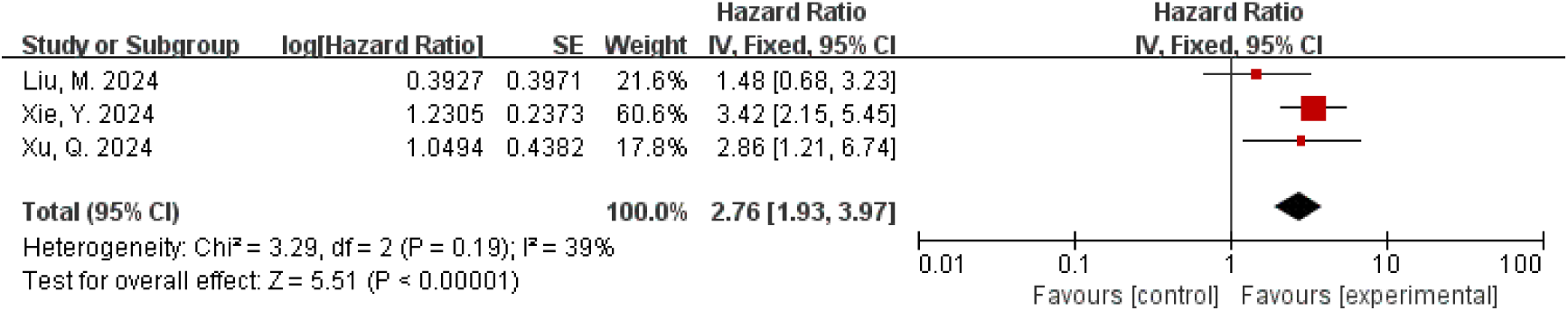
Forest plots of the relationship between FAI of LAD branch and MACE(FAI as a categorical variable)

### Correlation between FAI of LAD branch and MACE (FAI as a continuous variable)

The data is derived from 3^[6,36,40]^literature. The heterogeneity test resulted in chi^2^=0.37, DF=2(P= 0.83), and I^2^ = 0%, indicating a low level of heterogeneity. We employed fixed model meta analysis. The overall effect amount is as given in the figure: HR= 1.09, 95% CI= [1.01,1.16], P < 0.00001. There is a significant correlation between FAI of LAD branch and adverse cardiovascular events, according to the statistically significant results.(Figure 9)

**Figure 9.**
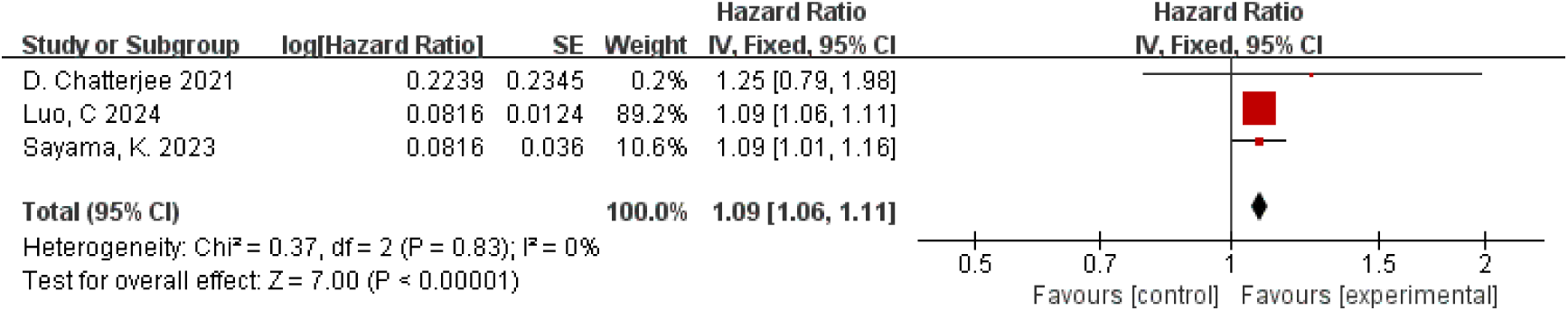
Forest plots of the relationship between FAI of LAD branch and MACE(FAI as a continuous variable)

### Correlation between FAI of LCx branch and MACE (FAI as a categorical variable)

The data is derived from 3^[35,43,44]^ literature. The heterogeneity test resulted in chi^2^=8.99, DF=2(P =0.01), and I^2^=78%, indicating a high level of heterogeneity. We employed random model meta analysis. The overall effect amount is as given in the figure: HR=2.68, 95% CI=[1.24, 5.80], P=0.01. There is a significant correlation between FAI of LCx branch and adverse cardiovascular events, according to the statistically significant results.(Figure 10)

**Figure 10.**
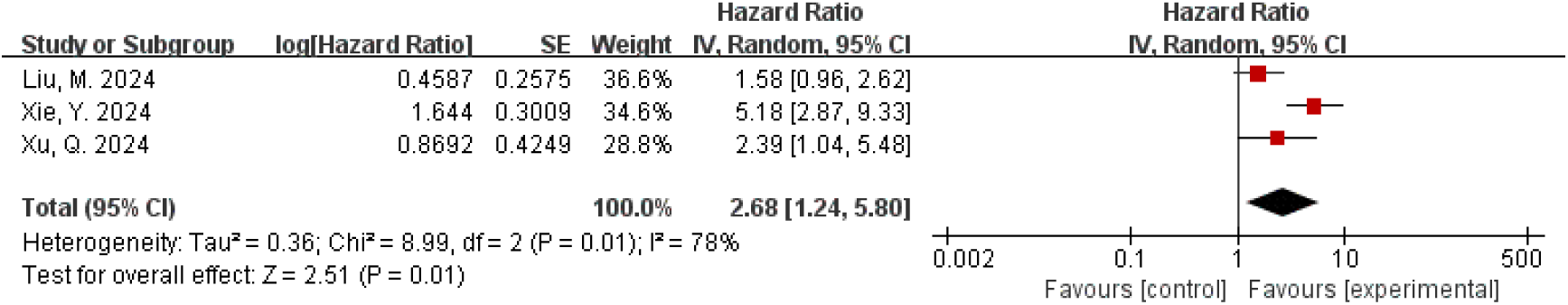
Forest plots of the relationship between FAI of LCx branch and MACE(FAI as a categorical variable)

### Correlation between FAI of LCX branch and MACE (FAI as a continuous variable)

The data is derived from 3^[6,36,40]^ literature. The heterogeneity test resulted in chi^2^=1.45, DF=2(P= 0.48), and I^2^=0%, indicating a low level of heterogeneity. We employed fixed model meta analysis. The overall effect amount is as given in the figure: HR=1.07, 95% CI = [1.05, 1.10], P < 0.00001. There is a significant correlation between FAI of LCX branch and adverse cardiovascular events, according to the statistically significant results.(Figure 11)

**Figure 11.**
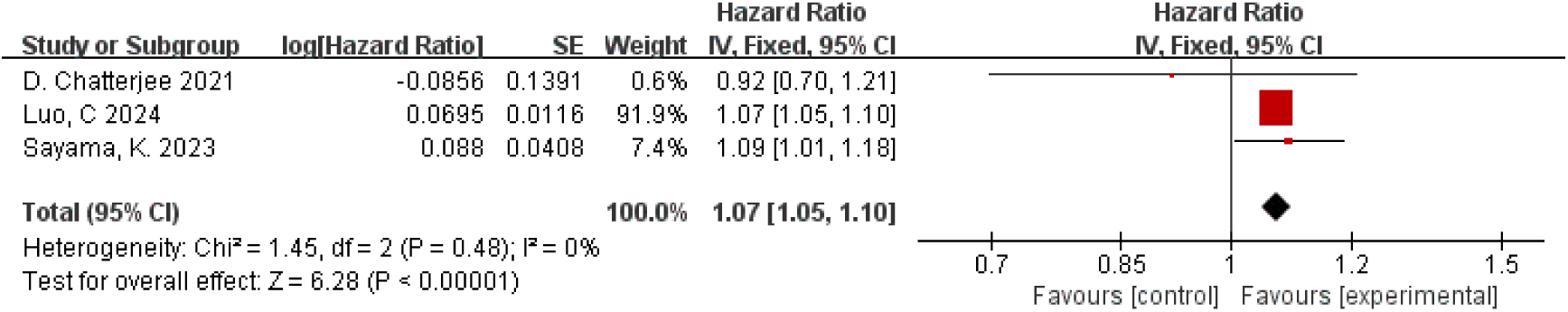
Forest plots of the relationship between FAI of LCX branch and MACE(FAI as a continuous variable)

## Discussion

The purpose of this meta-analysis is to investigate the relationship between FAI and adverse cardiovascular events. Our findings indicate a significant correlation between total FAI and branch FAI with adverse cardiovascular events,with those who have elevated FAI levels having a significantly higher risk of experiencing adverse cardiovascular events. This finding is in line with some findings from earlier studies^[48]^ and further supports the significance of FAI in assessing the risk of cardiovascular disease.

The fat attenuation index (FAI) derived from coronary computed tomography angiography (CCTA) is currently the most commonly used method to assess PCAT. The interaction between PCAT and coronary arteries provides the basis for its potential as a biomarker for cardiovascular disease.There is bidirectional biochemical communication between the coronary artery wall and PCAT. Under physiological conditions, PCAT secretes vascular protective factors, which have vasodilatory, anti-inflammatory, anti fibrotic, and antioxidant effects. Various bioactive factors act on vascular endothelial cells and vascular smooth muscle cells through paracrine or vascular secretion, maintaining cytokine balance and regulating inflammatory responses to external stimuli^[49]^.Under pathological conditions, after vascular endothelial cell injury, it triggers the “inside out” mode, inducing monocyte adhesion and lipid oxidation and releasing proinflammatory cytokines such as interleukin 6 (IL- 6), tumour necrosis factor alpha (TNF-α) and interferon gamma (IFN-γ). Stimulated PCAT also secretes cytokines such as IL-6 and TNF-α, which act on adjacent coronary arteries in a “paracrine” manner^[50]^, triggering the “outside-in” signalling mode, further exacerbating the inflammatory response of coronary arteries and promoting the development of atherosclerotic plaque, increasing the risk of adverse cardiac events^[51]^. There have been two similar meta-analyses in the past, and the scope of the included literature covers 2022^[52–53]^. However, the previous meta-analysis included relatively little literature and did not consider several recent emerging findings.

Compared with the previous meta-analysis, we believe that the results of the current meta-analysis on the association between FAI and MACE are the most recent.

However, this study also has some limitations. First, differences in statistical methods, diseases, study designs, regions, types of endpoints, etc. may potentially affect heterogeneity, resulting in high heterogeneity in this meta-analysis. Despite an extensive literature search, it is still difficult to completely exclude the possibility of publication bias, because studies with some negative results may not have been included due to their unpublished status, which could potentially affect the accuracy of the results. In addition, measurement methods and standards for FAI are not fully standardised and there is some heterogeneity in the definition of adverse cardiovascular events, which may affect the accuracy of the results. Meanwhile, the fact that most studies are observational makes it difficult to clarify the causal relationship between FAI and adverse cardiovascular events. More high-quality prospective and interventional studies are needed in the future to further validate this association.

The results of this study have important clinical implications, suggesting that clinicians should consider FAI as an indicator when assessing cardiovascular disease risk. Incorporating FAI into the cardiovascular disease risk assessment system may help to better identify high-risk populations and develop more targeted prevention and treatment strategies. For individuals with elevated FAI levels, lifestyle interventions such as a balanced diet and increased physical activity can be used to reduce FAI levels and thereby reduce the risk of cardiovascular disease. Looking ahead, more large-scale, multicentre, prospective studies are needed to further clarify the causal relationship and potential mechanisms between FAI and adverse cardiovascular events. On the other hand, efforts should be made to develop more accurate and convenient methods of FAI detection to promote their widespread use in clinical practice.

## Conclusion

Our preliminary evidence from the meta-analysis suggests a significant association between FAI and MACE, with individuals with elevated FAI levels having a significantly increased risk of developing MACE. However, due to potential biases in the included studies, the evidence may be limited and further evaluation is needed.

## Data Availability

All relevant data are within the manuscript and its Supporting Information files.

## Author contributions

**Conceptualization:** Yingzi Tan,Keji Chen.

**Data curation:**Yingzi Tan

**Formal analysis:** Yingzi Tan, Yaojian Wang, Boya Zhao.

**Methodology:** Yingzi Tan, Yuerong Jiang.

**Project administration:** Keji Chen.

**Supervision:** Yingzi Tan, Yuerong Jiang.

**Writing – initial draft:** Yingzi Tan.

**Writing – review & editing:** Yingzi Tan, Yuerong Jiang, Keji Chen.

Funding acquisition: Yuerong Jiang.

The research was financially supported by the funding from Major research project of scientific and technological innovation project of Chinese Academy of traditional Chinese Medicine (NO. CI2021A00908) and Hospital capability enhancement project of Xiyuan Hospital, CACMS. (NO. XYZX0101-14).

The authors have no conflict of interests to disclose.

